# Real World Performance of an Individualized Antitachycardia Pacing Algorithm

**DOI:** 10.1101/2024.12.31.24319831

**Authors:** Troy Jackson, Robert Taepke, Ulrika Birgersdotter-Green, Yong-Mei Cha, Jagmeet Singh, Alan Cheng, Raymond Yee

**Author notes:** **Corresponding Author:** Troy E. Jackson.

## Abstract

**Background:** A novel individualized antitachycardia pacing (IATP) algorithm using the post-pacing interval for real-time control has been introduced. Performance information is limited to a small safety and feasibility study with additional single-center and case studies. A larger-scale analysis is needed to better understand algorithm performance.

**Methods:** De-identified remote monitoring transmissions from devices with the IATP therapy applied were randomly selected. Rhythms were classified and effects of the novel algorithm were assessed. For monomorphic ventricular tachycardias (MVTs) proportions of successful therapy, shock-free episodes, and acceleration were calculated using Generalized Estimating Equations to correct for multiple episodes and compute statistics of the algorithm’s performance.

**Results:** There were 2259 MVT episodes in 336 patients. IATP succeeded in 87.1% of MVT episodes with 89.9% of MVT episodes ultimately free of shock therapy. Based on multivariate analysis, significant factors in therapy success were programming of at least the recommended number of sequences (90% at least recommended vs 73%, p=0.00088) and female sex (95% for female vs 86%, p=0.002). A trend to higher success was found for MVT with cycle length of 320ms or greater (90% vs. 83%, p=0.10). The IATP accelerated 3.6% of MVT episodes. None of the available factors was significantly associated with acceleration in the multivariate analysis.

**Conclusions:** The IATP algorithm succeeded in large proportion of MVT episodes and with low acceleration in patients randomly selected from remote monitoring transmissions. Using at least the recommended number of sequences had the strongest association with successful therapy.

**CLINICAL PERSPECTIVE:** *What is known:*

- A novel method for delivering antitachycardia pacing therapy using real-time feedback has been shown safe and feasible.
- Comparisons in computational studies and smaller case series suggest the novel method’s performance is superior to other antitachycardia pacing methods.

*What the study adds:*

- Under real-world programming and use, as evaluated on a large de-identified set of remote monitoring transmissions, the novel antitachycardia pacing algorithm treated monomorphic ventricular tachycardia episodes with high efficacy and a low proportion of acceleration.
- The strongest factor associated with successful therapy was programming at least the recommended number of pacing attempts.

## INTRODUCTION

The implantable cardioverter defibrillator (ICD) is a cornerstone strategy for mitigating the risk of death from sudden cardiac arrest.^1,2^ High-energy shock therapies are lifesaving but have deleterious effects on patients over time.^3^ Low energy antitachycardia pacing (ATP) is painless and has been shown effective in terminating monomorphic ventricular tachycardia (MVT) episodes, decreasing overall shock burden.^4,5,6^

In current practice, the standard ATP features programmed per consensus guidelines terminate roughly half of rapid VT episodes,^7,8,9^ leaving opportunity to improve patient experience^10^ and reduce healthcare utilization.^11^ In clinical study settings ATP can be tuned for higher efficacy,^4^ but this is difficult to achieve in real world settings even with concerted effort from clinicians.^12^ Additionally, the types of ventricular tachyarrhythmias seen in any given patient can vary, raising the need to adjust ATP programming for consistently effective therapy.

The Individualized ATP (IATP) feature is the first closed-loop ATP algorithm designed to adjust settings in real time to achieve VT termination success based on current patient conditions and arrhythmia characteristics.^13^ In brief, IATP is a burst plus extrastimulus protocol where the device automatically selects the number and timing of the stimuli and determines when to change to burst plus double extrastimuli by analyzing the post-pacing interval (PPI) in real time. There are two programmable parameters, the maximum number of attempts and the minimum extrastimulus interval.

Until now, information on the performance of IATP has been limited. The initial safety and feasibility study demonstrated successful algorithm operation and effectiveness in a highly selected population of ambulatory subjects.^13^ In a computational modeling comparison to burst ATP protocols, IATP improved efficacy.^14^ Multiple case studies have described unique aspects of IATP operation^15^ and termination of MVT refractory to burst and ramp ATP ^16,17,18,19^ while indicating that the problem of MVT acceleration remains.^20^ Most recently, a single-center comparison study found IATP effective both as a first therapy and, significantly, incrementally more effective after failed burst and ramp ATP.^21^ The study described here characterizes clinical outcomes of using IATP to terminate MVT in a real-world population with unconstrained device programming.

## METHODS

### Data Collection

This was a retrospective analysis using de-identified data obtained from a database which contains longitudinal stored device data collected by a remote monitoring system. The monitoring system included patients from the United States, Australia, Canada, and New Zealand. TJ had full access to all the data in the study and takes responsibility for its integrity and the data analysis.

The study population was selected from the database on May 16, 2022, and consisted of patients having ICD or Cardiac Resynchronization Therapy-Defibrillator (CRT-D) devices with IATP therapy enabled as the first therapy. Patient characteristics available in the database were limited to age at implant, sex, device type, and some attributes related to the indication for the device. Patients with at least one episode of attempted IATP therapy were randomly selected. Sampling was weighted to favor patients with multiple episodes so not more than about 50% of subjects had only a single episode. This was done to improve rhythm classification and to illuminate IATP performance on multiple MVTs. All IATP-treated episodes for a selected device were reviewed and classified for rhythm type and the effects of IATP therapy. Programming of parameters controlling tachyarrhythmia detection and IATP therapy at the time of the episode were extracted.

### Data Review and Endpoints

Each IATP-treated episode was reviewed by at least one author. Episodes with a ventricular rate similar to or slower than the atrial rate that lacked clear ventricular rhythm onset were reviewed by at least two authors. Reviewers classified the rhythm into which IATP was delivered and the effect of IATP (i.e. termination, acceleration, etc.).

Proportions of success and acceleration were calculated for all MVT episodes. Termination was defined as either a Type I break (at most 1 abnormal ventricular complex after ATP) or a Type II break (2 or more abnormal ventricular complexes after ATP that must differ in either cycle length or morphology compared to the pre-ATP rhythm). IATP Success was defined as an episode free of shock therapy and with termination by IATP, or spontaneous termination following an IATP attempt, or slowing of the MVT rate outside the programmed treatment rate following the IATP attempt. Shock-free success was defined similarly to IATP success, but with any type of ATP protocol comprising the final therapy of the episode, e.g., burst. Shock-free success takes into account IATP’s impact on the entire ATP strategy for the patient and allows comparisons to literature as studies often rely on device diagnostics to define success after classification of the index rhythm.^4,5,8^

Acceleration was classified by two criteria. Accelerated MVT was defined as a post-ATP MVT with a cycle length shorter than the pre-ATP cycle length by at least 30ms and at least 10% of the pre-ATP cycle length. Polymorphic acceleration was ATP converting MVT to polymorphic VT (PVT) or ventricular fibrillation (VF). Examples of efficacy and acceleration classifications are shown in Figure 1.

**Figure 1:**
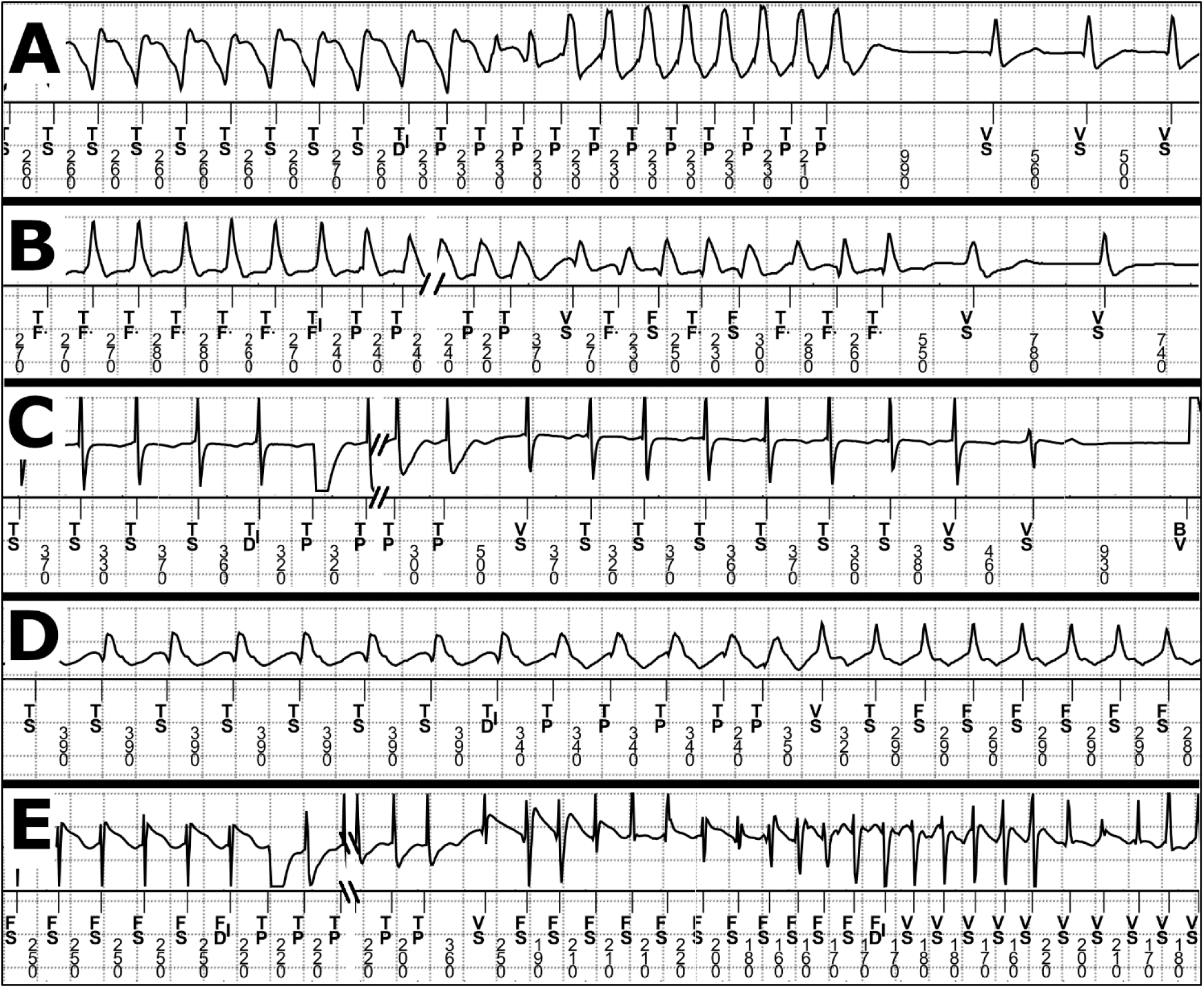
Definitions of ATP Effects with Examples. Each panel represents the possible effects of ATP therapy applied to monomorphic VT episodes. For each panel the top signal is a ventricular electrogram and the lower part is the event markers and the intervals between events in milliseconds. The broken timeline points marked with slashes are where ATP paces during the S1 portion of the train have been removed to allow better visibility of pre- and post-ATP rhythms. Panel A: Type I break, the ATP train is followed by no more than one abnormal ventricular event. Panel B: Type II break, the ATP train is followed by two or more abnormal events differing in cycle length or morphology from the rhythm prior to the ATP train before ultimately resolving to normal rhythm. Panel C: Spontaneous termination, the ATP train is followed by two or more abnormal beats that do not differ substantially in both morphology and cycle length from the pre-ATP rhythm prior to converting to normal rhythm. Panel D: Accelerated MVT, the post-ATP rhythm is monomorphic but has a cycle length shorter by at least 30ms and at least 10% of the pre-ATP rhythm. Panel E: Polymorphic acceleration, conversion of the rhythm to a polymorphic rhythm that sustains long enough to reach redetection. Event marker definitions are TP = ATP pace; BV = bi-ventricular pace; FS and TF. = detected intrinsic event with interval in the VF zone; TS = detected intrinsic event with interval in the VT zone; TD|, TF|, and TD| = point of sustained arrhythmia detection in the VT, FVT, and VF zones, respectively. VS = detected intrinsic event with interval outside of therapy zone or when rhythm is not being classified.

For investigating the effects of cycle length on IATP performance, *rapid VT* was defined as episodes with an initial cycle length of < 320ms (i.e., the nominal VF zone cutoff for the device). All other episodes were defined as *slower VT*. Episodes were also detected in device programmable zones, referred to by the abbreviations of VF, FVT, and VT. The FVT zone can be defined as FVT via VF or as FVT via VT, depending on whether it is programmed as a subzone of the VF or VT zone, respectively.

For evaluating the number of sequences to reach an effective ATP sequence the electrophysiologic efficacy was defined as the first ATP sequence to yield resumption of normal rhythm, even if VT reinitiated before the device identified episode termination using the criterion of eight consecutive intervals longer than any detection interval. In most cases an effective sequence ended the device episode.

Because of the variety of detection and therapy programming available, categorical values were assigned to ranges of the number of intervals to detect (NID) and number of ATP sequences programmed. Detection was classified as extended when the initial NID to detect VF was greater than 18/24 or the NID for VT was greater than 16; otherwise, detection was short. Similarly, in the device zone where the MVT was first detected, the programmed number of ATP sequences was categorized as either less than the nominal or at least nominal. Indication for the device was determined based on key terms in the device registration record. In the de-identification process, indication information entered by selecting “Other” and including free-text information was only represented as the keyword “Other”. Indications, where available, were assigned as secondary prevention, primary prevention, or other. The details of the terms and assignment algorithm are provided in the Supplemental Material.

### Statistical Analysis

Proportions of success and rhythm acceleration were computed for rhythms classified as monomorphic VT. Generalized estimating equations (GEE) were used to adjust event rates to account for patients with multiple episodes. Exploratory analyses of the frequencies of programmed detection and therapy configurations were evaluated as simple proportions. The impact of programming on the effects of IATP was determined using GEE analysis. Univariate and multivariate GEE analyses were performed to explore associations of successful ATP and acceleration with: age, sex, primary or secondary device indication, device type (ICD or CRT-D), de novo device or replacement, extended or short detection, at least nominal sequences or less than nominal, and rapid VT or slower VT. All data processing and analyses were performed using R Statistical Software (version 4.3.1; R Core Team 2023) with the gee package (gee: Generalized Estimation Equation Solver. version 4.13-26) and MATLAB with the Statistics and Machine Learning Toolbox (The MathWorks, Inc. Version 2024a).

## RESULTS

### Patient Characteristics

Prior to episode review, the weighted device sampling process resulted in 2601 episodes from 415 patients. Patient characteristics were available for 377 patients (90.8%) in the de-identified data set. After review there were 2259 episodes of MVT from 336 patients (81.0%). Patient characteristics for the entire sample and the subset who contributed at least one IATP-treated episode of MVT are summarized in Table 1. The distribution of the number of MVT episodes per patient is shown in Figure 2.

**Table 1:** Baseline Characteristics.

| **Baseline Characteristics** | **All Patients**<br>**(n = 415)** | **Patients with MVT**<br>**(n = 336)** |
| --- | --- | --- |
| **Demographic Data Available**<br>**(N, %)** | 377 (90.8%) | 307 (91.4%) |
| **Age (average years ± SD)** | 68.0 ± 13.3 | 68.9 ± 11.9 |
| **Male Sex (N, %)** | 308 (81.7%) | 260 (84.7%) |
| **First Device (N, %)** | 284 (75.3%) | 233 (75.9%) |
| **Indication: (N, %)** |  |  |
| **Secondary Prevention** | 160 (42.4%) | 138 (45.0%) |
| **Primary Prevention** | 203 (53.8%) | 156 (50.8%) |
| **Other/not specified** | 14 (3.7%) | 13 (4.2%) |
| **Device Type: (N, %)** |  |  |
| **Single-chamber ICD** | 65 (15.7%) | 42 (12.5%) |
| **Dual-chamber ICD** | 187 (45.1%) | 156 (46.4%) |
| **CRT-D** | 163 (39.2%) | 138 (41.1%) |

MVT, Monomorphic Ventricular Tachycardia; SD, Standard Deviation; ICD, Implantable
Cardioverter-Defibrillator; CRT-D, Cardiac Resynchronization Therapy - Defibrillator

**Figure 2:**
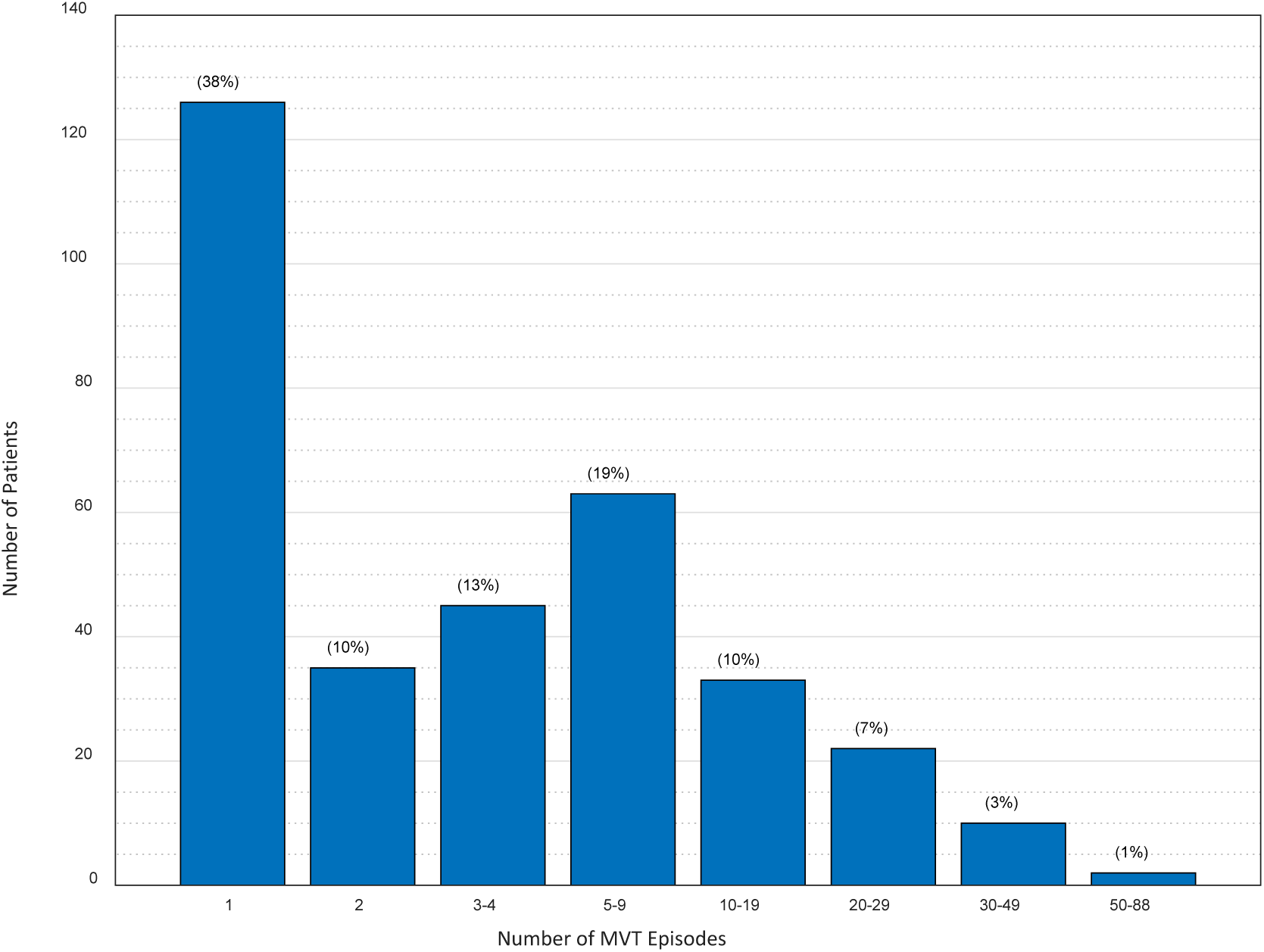
Frequency of MVT Episodes Per Patient. Histogram of the number of patients with different ranges of MVT episodes. The bins are sized in a semi-logarithmic progression with the percentage of the total patient group with MVT noted at the top of each bar.

### Episode Characteristics

MVT was the rhythm classification at initial detection in 2259/2601 episodes (86.8%). The median cycle length of MVT was 340 ms with 30.8% of episodes presenting as rapid VT. The full distribution of MVT cycle lengths is provided in Figure 3. The most common non-MVT rhythms were 228 (8.8%) supraventricular tachycardia (SVT) episodes, followed by 94 (3.6%) episodes of polymorphic ventricular tachycardia (PVT) or VF. Least common were 19 (0.7%) non-sustained VT (NSVT) episodes, and one (0.04%) T-wave oversensing (TWOS) episode.

**Figure 3:**
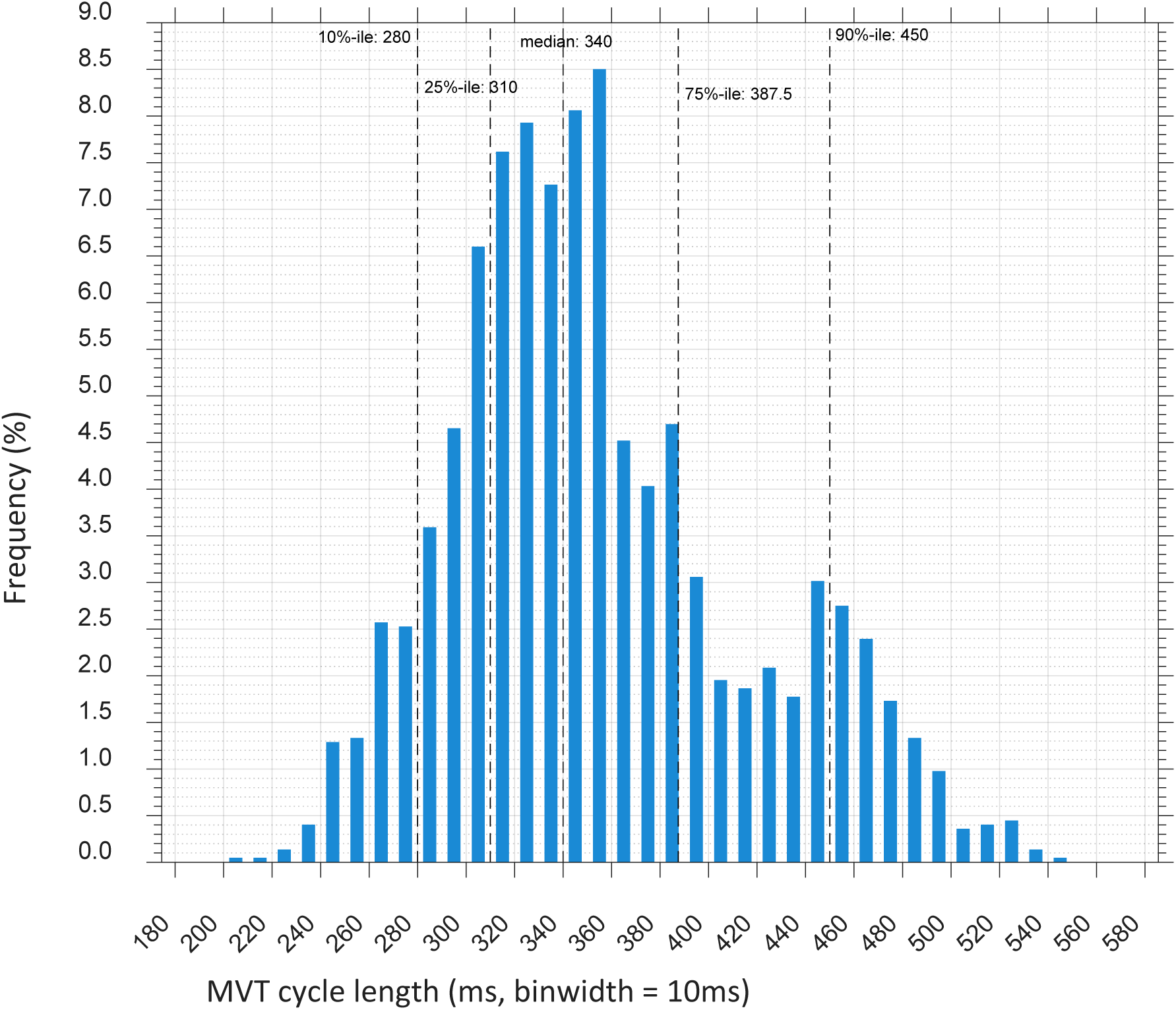
Cycle Length at Initial Detection for MVT Episodes. Histogram of the cycle length calculated at the first detection and IATP application with summary percentiles noted in milliseconds. %-ile = percentile.

### Success of IATP and overall shock-free success

Outcomes of episodes of MVT with IATP therapy are summarized in Figure 4. The GEE-adjusted IATP success was 87.1% (95% CI 84.4% to 89.5%). The GEE-adjusted shock-free proportion of all ATP was 89.9% (95% CI 87.6% to 91.8%). Univariate analysis of the effect of VT cycle length resulted in a GEE-adjusted IATP success for rapid VT of 82.7% (95% CI 78.2% to 86.4%) vs 90.5% for slower VT (95% CI 87.7% to 92.7%), a significant difference (p = 0.00006). Likewise, univariate GEE-adjusted shock-free proportion of rapid VT was 86.1% (95% CI 82.4% to 89.2%) vs 92.6% (95% CI 90.2% to 94.4%) also significantly different (p = 0.0002).

**Figure 4:**
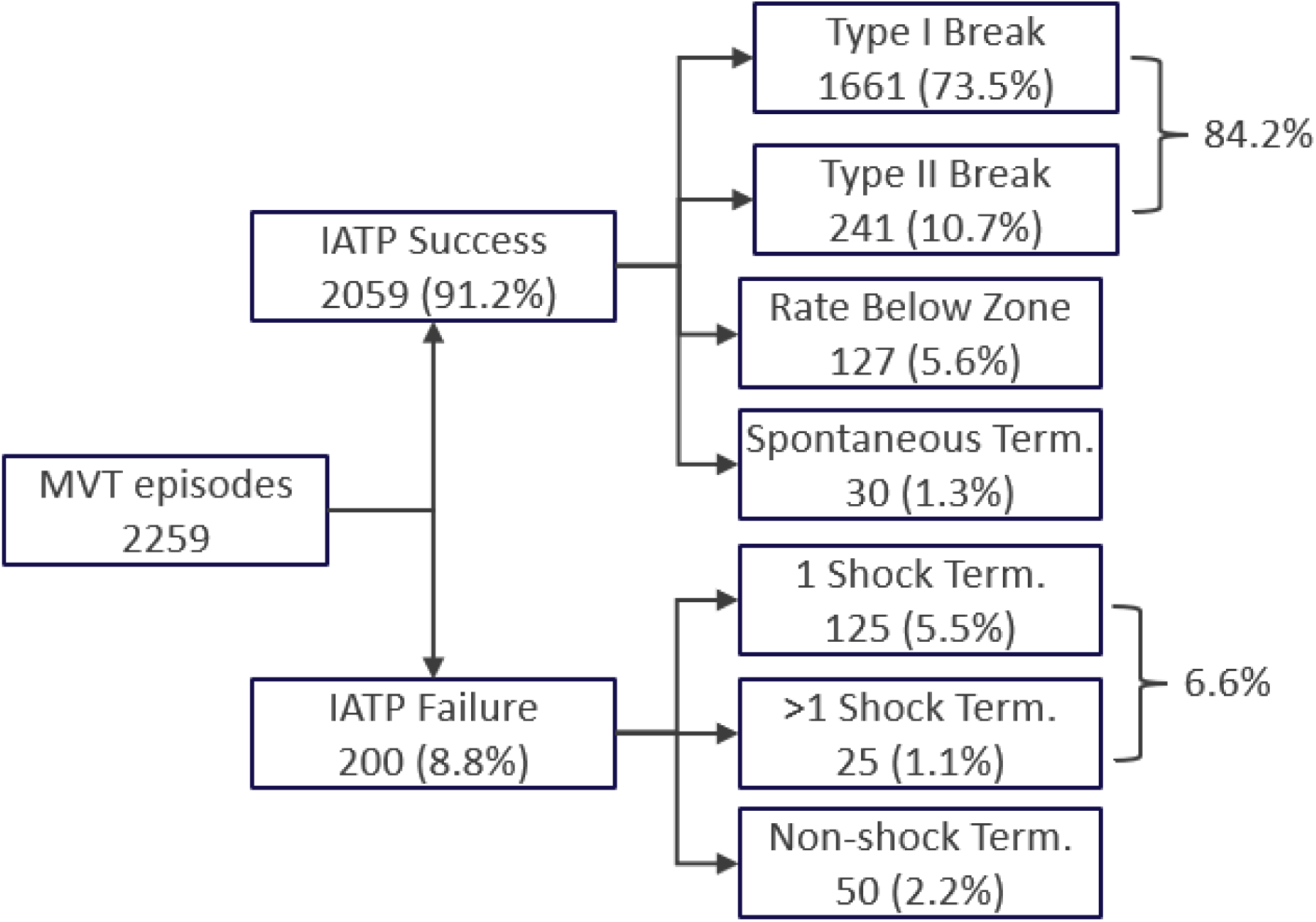
Outcomes of IATP-treated MVT Episodes. The final outcome of each MVT episode treated by IATP. Percentages are of the total number of MVT episodes. “Rate below zone” indicates that a tachyarrhythmia was present at the end of the episode but was below the programmed therapy rates. Term. = Termination.

From a patient-level perspective, IATP was successful for at least one MVT episode in 92% of patients, at least one rapid VT episode in 85% of patients with rapid VT, and at least one slower VT episode in 95% of patients with slower VT. In 234/336 patients (70%) IATP was successful for all MVT episodes. The number of MVT episodes per subject in this always-successful group ranged from one to 88, with a median of 2 episodes. In 26/336 patients (7.7%) IATP failed for all MVT episodes, in this group the number of episodes per subject ranged from 1 to 8, with a median of 1 episode.

Figure 5 illustrates the per-sequence and cumulative electrophysiologic efficacy for IATP. The first sequence was effective in 70.6% of the episodes (top of first stacked bar), but in 4.4% MVT recurred prior to device recognition of rhythm termination, resulting in first sequence IATP success of 66.2%, GEE-adjusted to 66.9% (95% CI 63.0% to 70.6%). By design the IATP algorithm always starts with a single extrastimulus (S2) and can apply up to two extrastimuli (S2/S3). The earliest S3 may be applied is the third sequence. An S3 was applied in 115 MVT episodes (5.2%) in 51 patients. The proportion of sequences that included an S3 increased progressively from the third (2%) to the seventh (69%) sequence although the absolute number of episodes receiving a high number of sequences was small. Addition of S3 resulted in higher effectiveness for episodes with more sequences (see the bottom row in Figure 5 for the 6^th^ and 7^th^ sequences). The cumulative effectiveness of double extrastimuli in terminating both the MVT and the device episode was 40%, a GEE-adjusted rate of 37% (95% CI 27% to 49%).

**Figure 5:**
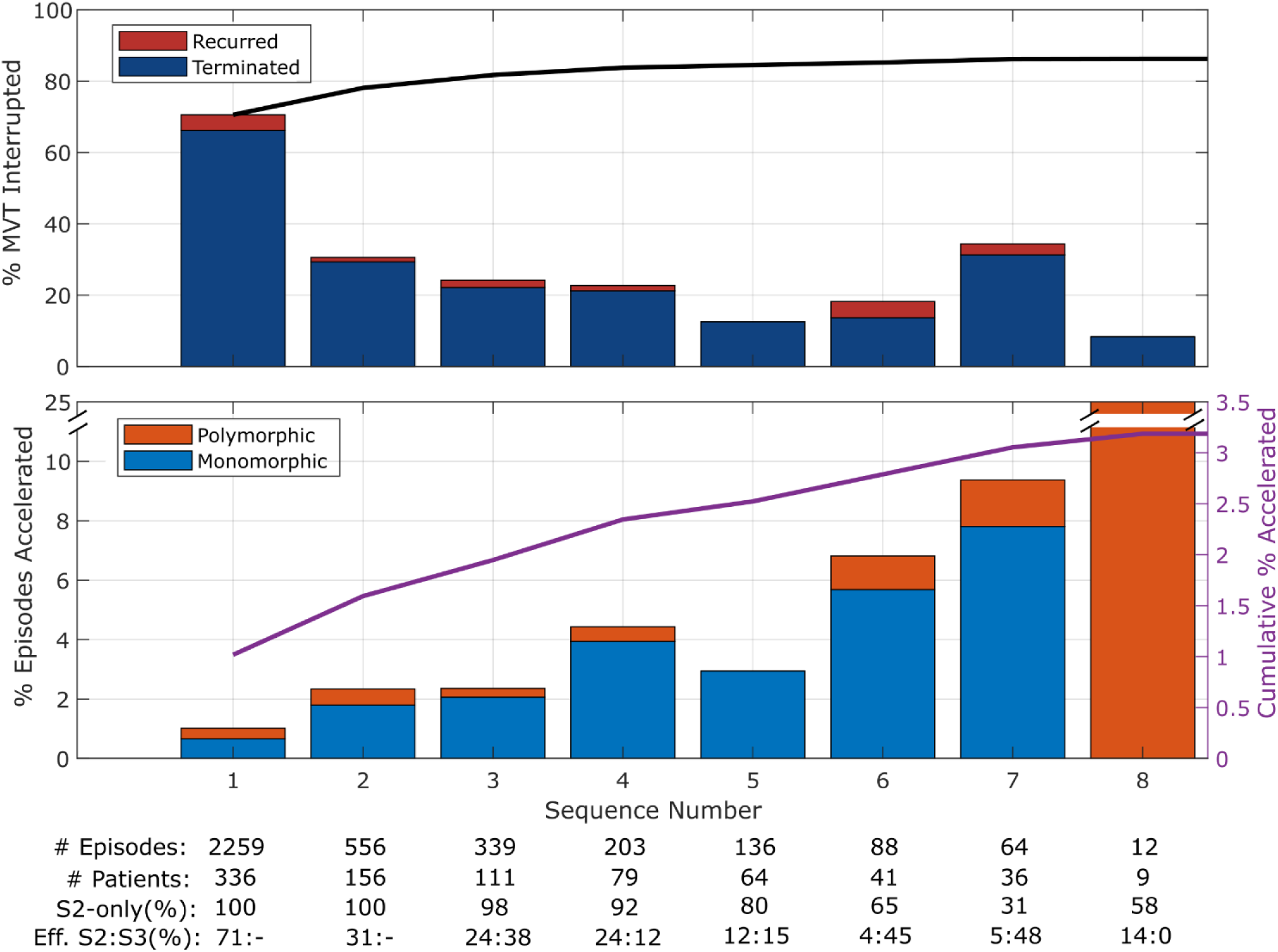
Effects of IATP by Sequence. The effects of IATP on MVT episodes at each sequence. The upper graph shows the effect of each sequence until electrophysiologic and device-defined episode termination by IATP or until the first restoration of normal rhythm with MVT recurring before the device termination criteria were satisfied. The cumulative rate of interruption of the MVT is shown in the black line. The lower graph is the acceleration rate for each sequence indicating if the accelerated rhythm was monomorphic or polymorphic. The purple line and right vertical axis are the cumulative acceleration rate. Note the small number of episodes at sequence 8 produces an outsized proportion of acceleration (25%, 3 episodes of 12) and is truncated on the plot. Not shown are the outcomes of the 9 episodes that had more than 8 sequences, one successfully terminated on the 12^th^ sequence and none of the 9 accelerated. S2-only = percentage of sequences where a single extrastimulus was used. Effective S2:S3 = the efficacy for each type of sequence applied, e.g. 24:38 is a 24% effective proportion of treatment by S2-only, and 38% effective treatment by sequences with an added S3.

The potential importance of multi-sequence IATP in reducing shock therapy for a patient was estimated by determining for each patient the largest number of sequences for a successful IATP treated episode. The proportions of patients at each maximum sequence value were calculated over the entire patient cohort and in the subgroup of patients were IATP was always successful. Nine percent of the always-successful patient group had at least one episode where more than four sequences were delivered (see Figure 6). For the entire set of patients, 13% had at least one successful episode with more than four sequences delivered.

**Figure 6:**
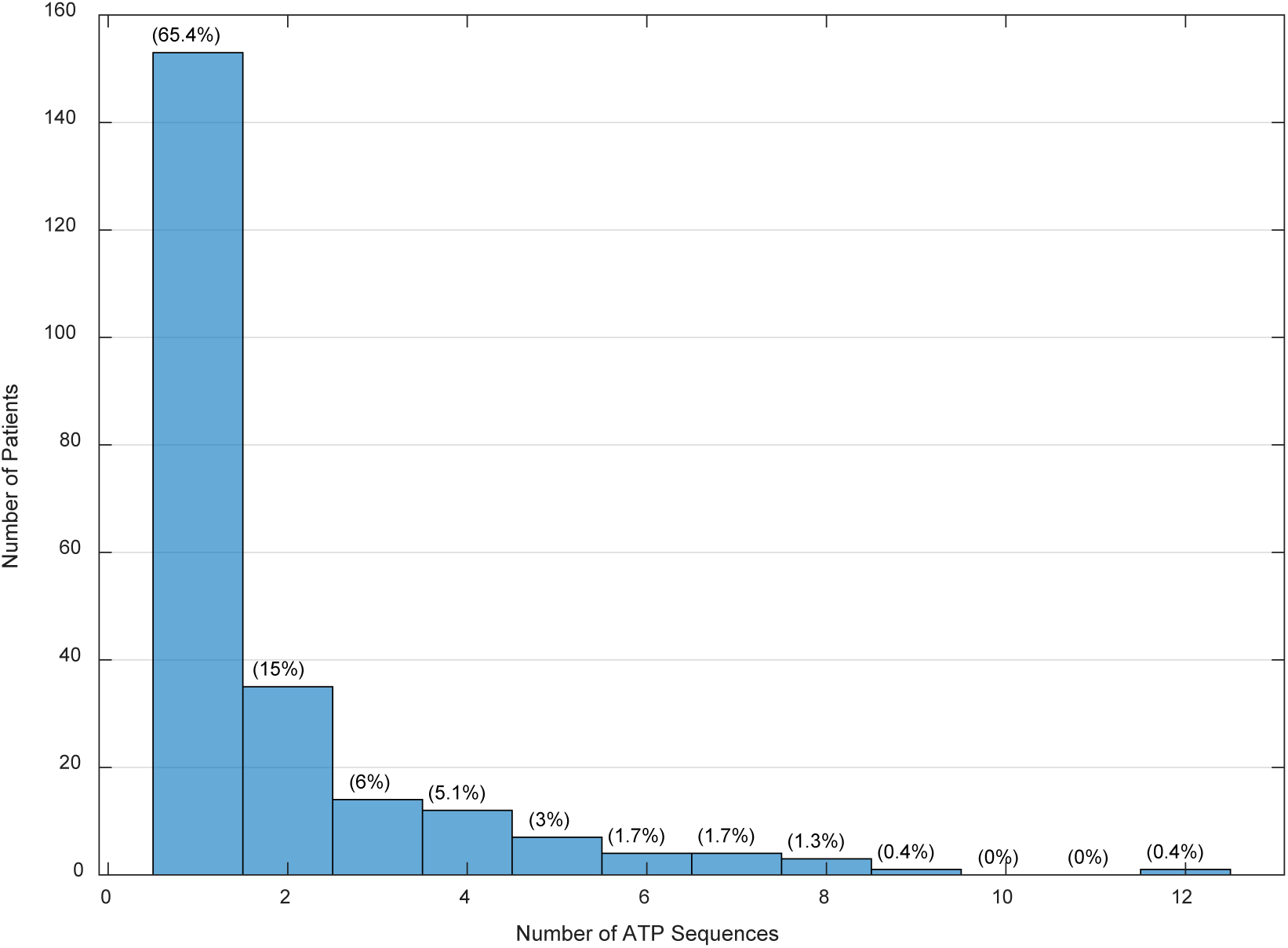
Sequences required in patients with always-successful IATP. For patients in which IATP was always successful, the maximum number of ATP sequences applied for any MVT episode is the horizontal axis. Each bar shows the number and percentage of patients in the always-successful group needing that maximum. The maximum number of sequences that can be programmed for a single therapy is 10, the patient with 12 had IATP set as the first two therapies.

### Tachycardia Detection and Therapy Programming

Because all therapy and detection programming was at physician discretion and could change over time, we characterized programming practices. Programmed detection settings for the zone at initial detection of the MVT were available for all episodes. Therapy settings could not be retrieved for eleven episodes among six patients. With unconstrained programming there was a high degree of variation among patients, the details of which are provided in the Supplemental Material.

For 268 patients with at least one episode of MVT in the VT zone (including FVT via VT), the median programmed zone cutoff was 380ms with a range of 300ms to 600ms. The initial number of intervals to detect (NID) was most commonly the nominal value of 16 (48% of patients). The median NID was 20 with a range from 12 to 100 intervals.

There were 99 patients with at least one episode of MVT in the VF zone (including FVT via VF). The median programmed VF zone cutoff was 300ms with a range of 250ms to 360ms. The initial number of intervals to detect was frequently the nominal value of 30/40 (81% of patients). The median NID was 30/40 with a range from 12/16 to 45/60 intervals.

Two parameters are used to configure IATP therapy in each zone: the maximum number of sequences to attempt, and the minimum extrastimulus interval (Minimum S2/S3). The Minimum S2/S3 was the nominal value for the applicable zone in more than 90% of patients (details in Supplemental Table S3). The maximum number of sequences was programmed to at least the nominal value for the respective zone in 55%, 76%, and 76% of patients for the VF, FVT, and VT zones, respectively. Complete information on the number of sequences programmed is provided in Table 2. More detailed information about detection and therapy programming, including the use of the FVT subzone, is provided in the Supplemental Material.

**Table 2:** Programming of Number of Sequences for Patients with Treated MVT in a zone.

| **Number of Sequences** | **VF Zone (70 configurations)** | **FVT zone (47 configurations)** | **VT Zone (280 configurations)** |
| --- | --- | --- | --- |
| **1** |  | 0% | 0% |
| **2** | 43% | 0% | 1% |
| **3** | 56% (nominal) | 15% | 5% |
| **4** | 1% | 2% | 2% |
| **5** |  | 76% (nominal) | 1% |
| **6** |  | 0% | 2% |
| **7** |  | 2% | 76% (nominal) |
| **8** |  | 2% | 4% |
| **9** |  | 0% | 0.4% |
| **10 (maximum)** |  | 2% | 8% |

VF, Ventricular Fibrillation; FVT, Fast Ventricular Tachycardia; VT, Ventricular Tachycardia

### Factors associated with successful IATP

Both univariate and multivariate GEE analyses were performed to determine if any factors were associated with successful IATP. In reviewing episode details, there were no episodes where the programmed value of Minimum S2/S3 influenced the delivery of IATP and the parameter was not included in the analysis. The two factors with significant association with successful IATP in the multivariate analysis were number of sequences programmed and sex (see Table 3).

**Table 3:**
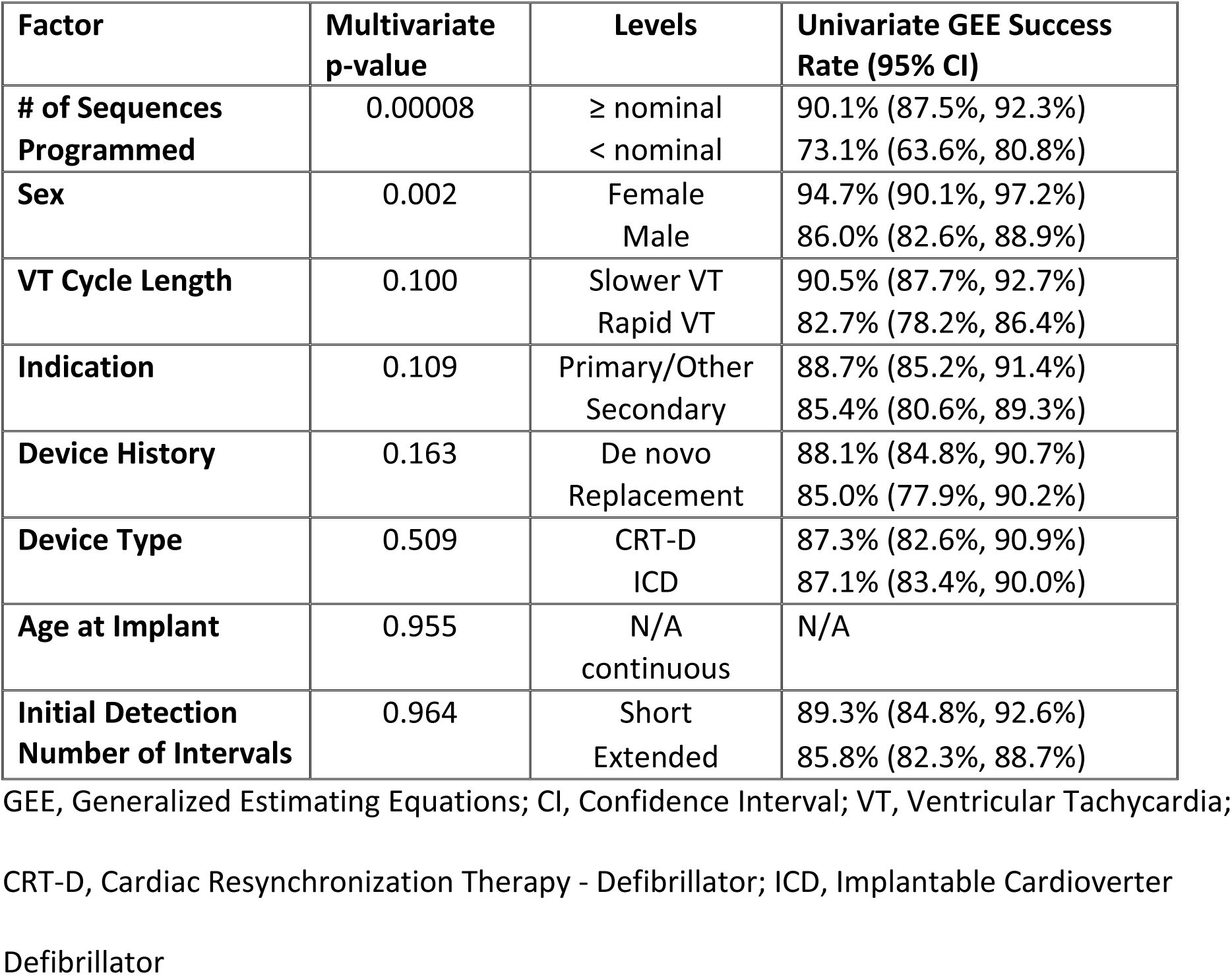
Analysis of Factors for Association with Successful IATP.

| **Factor** | **Multivariate**<br>**p-value** | **Levels** | **Univariate GEE Success**<br>**Rate (95% CI)** |
| --- | --- | --- | --- |
| **# of Sequences**<br>**Programmed** | 0.00008 | ≥ nominal<br>< nominal | 90.1% (87.5%, 92.3%)<br>73.1% (63.6%, 80.8%) |
| **Sex** | 0.002 | Female<br>Male | 94.7% (90.1%, 97.2%)<br>86.0% (82.6%, 88.9%) |
| **VT Cycle Length** | 0.100 | Slower VT<br>Rapid VT | 90.5% (87.7%, 92.7%)<br>82.7% (78.2%, 86.4%) |
| **Indication** | 0.109 | Primary/Other<br>Secondary | 88.7% (85.2%, 91.4%)<br>85.4% (80.6%, 89.3%) |
| **Device History** | 0.163 | De novo<br>Replacement | 88.1% (84.8%, 90.7%)<br>85.0% (77.9%, 90.2%) |
| **Device Type** | 0.509 | CRT-D<br>ICD | 87.3% (82.6%, 90.9%)<br>87.1% (83.4%, 90.0%) |
| **Age at Implant** | 0.955 | N/A<br>continuous | N/A |
| **Initial Detection**<br>**Number of Intervals** | 0.964 | Short<br>Extended | 89.3% (84.8%, 92.6%)<br>85.8% (82.3%, 88.7%) |

GEE, Generalized Estimating Equations; CI, Confidence Interval; VT, Ventricular Tachycardia;
CRT-D, Cardiac Resynchronization Therapy - Defibrillator; ICD, Implantable Cardioverter
Defibrillator

### Safety of IATP

#### Acceleration of MVT by IATP

Overall acceleration was low, with 2.9% of the episodes accelerated, 0.7% to PVT/VF and 2.2% remaining MVT. The GEE-adjusted overall acceleration rate was 3.6% with adjusted rates for PVT/VF and accelerated MVT of 0.9% and 2.6% respectively. The outcomes of episodes accelerated by IATP are summarized in Figure 7, and the per-sequence acceleration rate is shown in Figure 5. None of the available factors reached significance in the multivariate analysis for overall acceleration rate, rate of accelerated MVT, and rate of acceleration to PVT/VF.

**Figure 7:**
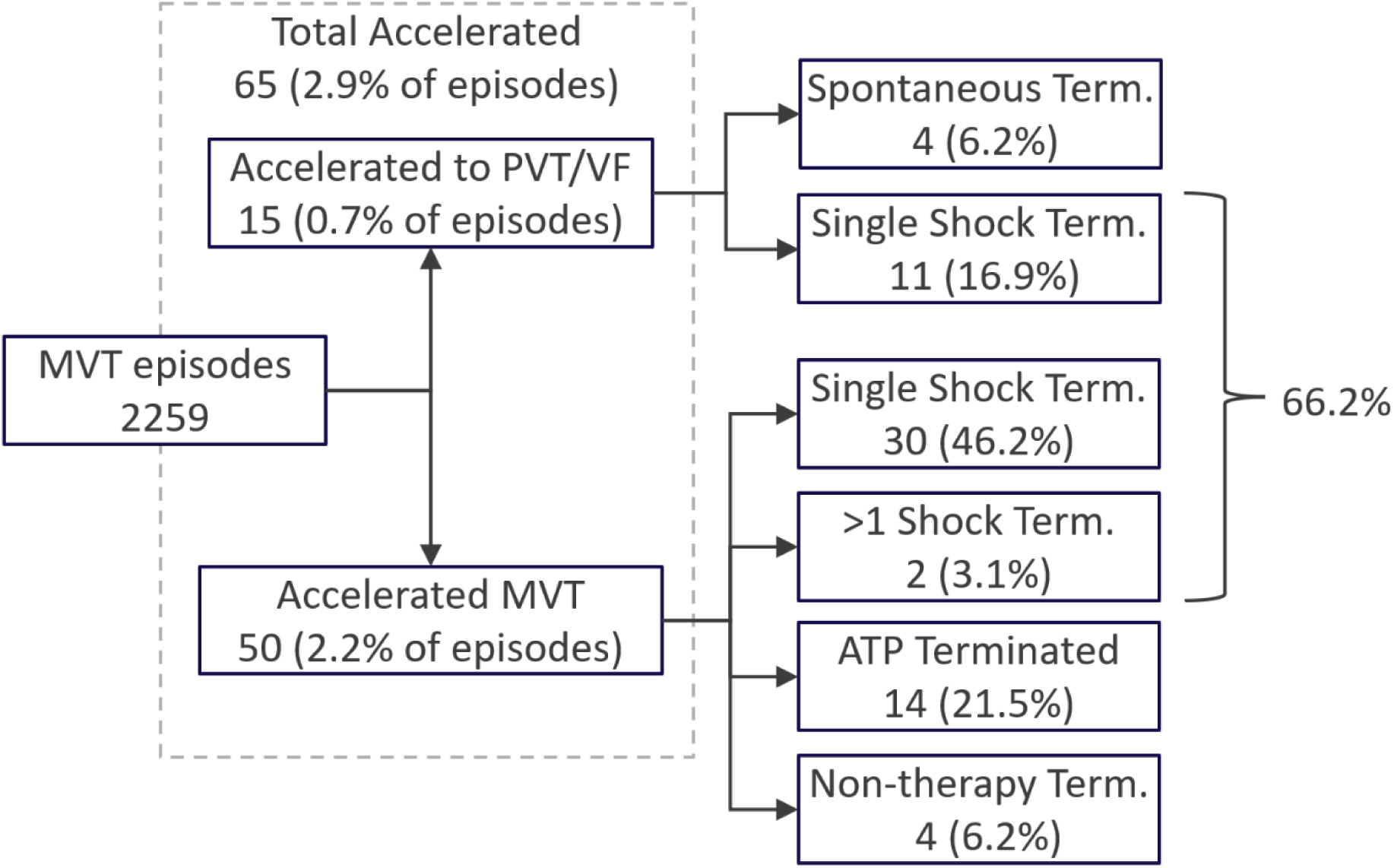
Summary of Episodes Accelerated by IATP. The outcome of all episodes accelerated by IATP. Percentages are of the total number of accelerated episodes, unless noted otherwise. Term. = Termination.

When evaluating differences between rapid VT and slower VT episodes, there was a significantly higher proportion of FVT episodes accelerated to PVT/VF (1.8% vs 0.4%, P = 0.004), but this association did not reach significance in the multivariate analysis (P = 0.10). The overall acceleration rate of 4.3% for rapid VT and 3.1% for slower VT was not significantly different, nor were the accelerated MVT rates of 2.5% and 2.6% for rapid VT and slower VT, respectively.

#### Safety of Inappropriately Delivered IATP

The most frequent inappropriately treated rhythm was SVT. There were 247 SVT episodes treated by IATP. De novo SVT was encountered 228 times (92%) while SVT was treated in 19 episodes when IATP terminated MVT occurring during SVT, but the SVT persisted at a therapy-eligible rate and further IATP therapy was applied. In total there were 643 sequences of IATP delivered into SVT in 86 patients. Pro-arrhythmic effects were observed in 33 sequences from 30 episodes in 13 patients. Forty-eight percent of those sequences produced single or double PVCs. There were 10 sequences that induced non-sustained VT in 10 episodes from 5 patients. Sustained MVT was induced in 5 episodes (2.0%) from 4 patients (4.6%), three episodes of which were from those where MVT termination occurred during SVT. All induced MVT was converted by subsequent therapy, ATP in three episodes, and shock in two episodes. In two patients, each with a single episode of SVT, the first sequence of IATP induced VF which was converted by a single shock. The overall rate of induction of sustained MVT and VF was 2.8% and induction of sustained ventricular arrhythmia requiring shock therapy was 1.6%.

There were 19 NSVT episodes and one TWOS episode with applied IATP. There were no induced ventricular arrhythmias. IATP sequences either ended the episode (11) or had no effect (19). Episodes either resolved spontaneously (15) or ended with shock therapy (5).

## DISCUSSION

This analysis of an Individualized ATP algorithm via de-identified device transmissions represents the first large-scale performance report on the algorithm in the setting of real-world programming of devices. The IATP algorithm was highly effective for MVT with an overall success of 87% and a 90% shock-free proportion. Significant associations with successful IATP in the multivariate model were found for female sex and use of at least nominal number of sequences; but not for MVT cycle length (<320ms vs ≥ 320ms), device type (do novo vs replacement and ICD vs CRT-D), age at implant, secondary prevention indication, and short vs extended detection durations. The overall acceleration rate of 3.6% was not significantly associated with any factors in multivariate analysis.

Comparing performance of ATP across different reports in the literature is inherently limited by the impacts of different detection and therapy settings, different methods for defining successful ATP, the size of the population analyzed, and whether statistical adjustments are applied for multiple episodes from the same patient. The post-hoc analysis of ATP effectiveness from PainFREE SST (PF SST) is the closest match to the present study considering the population size, statistical methodology (applying GEE-adjustment and using the same definition of shock-free success), proportions of secondary prevention, CRT-D devices, programming of extended detection duration, and the distribution of VTCL.^4^ Shock-free success in the IATP cohort was 89.9% vs 81.4% for PF SST. Both studies defined rapid VT as VTCL < 320ms, with IATP success of 82.7% vs 75.0% success for PF SST. Higher success was also seen for slower VT with IATP successful in 90.5% of episodes vs 88.0% for PF SST. To provide further context, a single-center retrospective study on patients programmed to the center’s approach of “progressive ATP and shock therapy” for VT in the range of 150 bpm to 230 bpm reported success rates from 88% to 91% when programming 6 ATP attempts.^22^ That success rate is comparable to the 88% for all zones in the IATP cohort.

The impact of multiple ATP attempts in an episode was also reported for PF SST. Both studies demonstrated that across VT cycle length ranges, additional attempts increase effectiveness. Both analyses show diminishing returns to the cumulative episode success rate after the third or fourth sequence. However, Figure 6, demonstrates that 9% of 234 patients with completely successful IATP therapy required more than four sequences in at least one episode. Extending to all 336 IATP treated patients showed 13% of patients had a successful episode with more than four sequences applied. Martins, et. al. reported a similar finding in their single-center evaluation of programming ten sequences of ATP for rapid MVT.^23^ Thus, focusing on cumulative efficacy can mislead about the necessary number of sequences needed to minimize shock therapy. Similarly, a retrospective analysis of the UMBRELLA Registry, showed that using more than one ATP attempt for rapid VT reduced the risk of receiving a shock by 52% compared to single attempt.^24^ Although in both the UMBRELLA and Martins studies detection durations were shorter than current guidance, potentially exaggerating the effect.

For prospective trials in the delayed therapy era, the focus has been on rapid VT (>188bpm or >200 bpm) and a single burst ATP attempt at 88% VTCL. Two studies reported GEE-adjusted efficacy of 46.9%^9^ and 49%^25^ and a third reported unadjusted efficacy of 52%.^8^ These are lower than the GEE-adjusted IATP first sequence success rate of 66.9%, hypothetically because the consistent application of S2 less than 88% VTCL on the first sequence yields higher initial sequence success.

The role of programming in IATP effectiveness in these analyses clearly illustrated a strong association between the use of at least the nominal sequence programming and IATP success rate. The nominal values for number of sequences were based on an analysis of the safety and feasibility study.^13^ As IATP does not use a second extrastimulus until a single extrastimulus approach becomes futile upon loss of S2 capture, the nominal sequence recommendations were set to increase the probability that an S3 would be introduced before the sequence limit was reached.

There has been one prospective evaluation of combining IATP and conventional ATP (burst or burst then ramp), with the order of therapy assigned sequentially.^21^ The success rates were not adjusted for multiple episodes per patient and there were 11 treated patients in each group. The success of conventional ATP was lower as the first therapy (80% vs IATP’s 85%) and as the final therapy (89% vs IATP’s 93%), with neither difference reaching statistical significance. It was noted that IATP was more successful in converting episodes in which prior conventional ATP failed, converting all 6 episodes in 5 patients in which IATP could be applied while conventional ATP converted only 3 of 9 episodes from 3 patients when prior IATP failed. The difference was significant (p = 0.028) and hints at different interactions with VT between burst plus extrastimulus and other ATP protocols.

In looking at the safety aspects of ATP therapy we addressed acceleration and the effects of delivering IATP inappropriately for detections of VT/VF for SVT, NSVT, and TWOS. In the literature acceleration rates are rarely GEE-adjusted, with only Strik, et al. reporting a GEE-adjusted acceleration rate of 6.6% cumulative for up to 6 bursts, comparable to the IATP overall rate of 3.6%.^22^ In more recent single burst ATP studies, reported unadjusted acceleration rates for rapid VT were 9.4%^9^ and 15.2%.^8^ These are higher than the 4.4% acceleration rate for rapid VT in the IATP cohort, which were always programmed to at least two ATP attempts.

Interestingly, while the rate of rapid VT acceleration to polymorphic rhythm for IATP was 1.8% (GEE-adjusted) and 0.9% in PRAETORIAN^9^, Strik et, al. reported that over half of the accelerations in their analysis were to VF.^22^ The difference with Strik may indicate a mechanistic difference when applying multiple attempts between burst and ramp compared to the burst plus extrastimulus of IATP, as IATP decreases only the extrastimulus below 88% of VTCL.

There is limited high-quality evidence concerning safety of ATP for SVT or oversensing. The Italian Clinical Service Project reported 353 inappropriate ATP deliveries for atrial fibrillation in a CRT-D population.^26^ Sustained ventricular tachyarrhythmia was induced only in 0.3% (upper 95% confidence bound of 1.6%). A retrospective analysis of the OMNI Trial found that ATP delivered into 176 episodes of SVT resulted induced ventricular arrhythmias in 3.4% (95% CI 1.3% to 7.3%).^27^ In the present study, IATP into SVT induced sustained ventricular arrhythmia in 2.8 % of episodes. Thus, it appears that IATP is unlikely to be more pro-arrhythmic than other ATP protocols when delivered inappropriately.

### Limitations

As a retrospective analysis without a contemporary control group, comparisons in these analyses are directional, and cannot be quantitative because of variations in patient populations, therapy strategies, and analysis methods. Devices capable of IATP come from a single manufacturer and single model family which introduces selection bias for both choice of device and the decision to choose IATP as a therapy. The limited demographic data available meant the treated patient population is not well defined, however the available data were typical of large, general indication ICD/CRT-D trials. Device programming was not standardized and left to physician judgment and could be amended any time. Finally, the analysis was limited to remote transmissions, so the clinical consequences of episodes are unknown.

## Conclusion

The Individualized ATP feature provided high rates of ATP success in the treatment of monomorphic VT across the spectrum of VT cycle lengths and in most patients who were randomly selected from remote monitoring transmissions. Acceleration was low and most accelerated rhythms remained monomorphic VT. Use of at least the nominal number of sequences was strongly associated with successful IATP.

## Data Availability

The data that support the findings of this study are available from the corresponding author upon reasonable request.

## Acknowledgements

We would like to thank Paul Belk for his foundational contributions to the design and study of IATP and Recce Holbrook for assisting in the preparation of this manuscript.

## Supplemental Material

Assignment of Indication for Device

VT and VF Zone Detection Programming

FVT Sub-Zone Programming

Therapy Programming

Figures S1-S4

Tables S1-S3

## Non-standard Abbreviations and Acronyms

ATP: antitachycardia pacing
CI: confidence interval
CRT-D: cardiac resynchronization therapy-defibrillator
FVT: fast ventricular tachycardia
GEE: generalized estimating equations
ICD: implantable cardioverter-defibrillator
MVT: monomorphic ventricular tachycardia
NID: number of intervals to detect
NSVT: non-sustained ventricular tachycardia
PPI: post-pacing interval
PVT: polymorphic ventricular tachycardia
S1: initial constant cycle length stimulus train
S2: first extrastimulus
S3: second extrastimulus
SVT: supraventricular tachycardia
TWOS: T-wave over-sensing
VF: ventricular fibrillation
VT: ventricular tachycardia

## Supplemental Material

### Supplemental Material: Real World Performance of an Individualized Antitachycardia Pacing Algorithm (IATP)

#### Assignment of Indication for Device

For devices with available demographic data, an indication for the device was assigned based on a hierarchical algorithm. Key terms identifying secondary prevention criteria were given highest priority, followed by specific primary prevention terms, then inferences from more general terms. The specific terms are provided in Table S1.

#### VT and VF Zone Detection Programming

The span of VT detection intervals was broad, ranging from 300ms to 600ms and is detailed in Figure S1. Changes to a longer VT zone cutoff were observed in 35 patients (13%). The number of intervals to detect VT also spanned the minimum and maximum programmable values in the device. The guideline directed value of 24 was used in 18% of programming configurations. The contribution of MVT episodes for each VT NID value is provided in Figure S2. Changes to a larger NID were seen in 14 patients.

The detection programming of the VF zone was less varied than the VT zone. The longest VF zone cutoff was 360 ms and the shortest was 250 ms. The most common VF zone cutoffs were 300ms and 320ms.

The full distribution of VF zone programming is provided in Figure S3. There were two patients with a change to a longer VF zone cutoff. For the number of intervals to detect, the nominal value of 30/40 intervals dominated, with 80% of configurations set to that value. There were no changes to VF NID. Despite guideline recommendations there were some shorter NIDs of 18/24 and 12/16. The number of episodes detected at each value of VF NID is shown in Figure S4.

#### FVT Sub-zone Programming

There were 46 patients that had 139 episodes of MVT occur in a Fast VT sub-zone, the majority of which (103 episodes from 34 patients) were in the FVT via VF configuration. The FVT via VT configuration was only used in seven patients, with 19 episodes of MVT. The FVT zone cutoffs used are provided in Table S2.

#### Therapy Programming

For the VF zone, only a single ATP therapy can be selected, so when IATP was applied in the VF zone it was as the first and only ATP therapy in the zone. For the FVT and VT zones, IATP could also be used as a later therapy. In the FVT zone, one patient also had IATP as the second therapy which was applied in one episode. In the VT zone, first and second therapy set as IATP was found in 19 episodes from 10 patients. A single patient had IATP set to the first three therapies in the VT zone in 30 episodes.

The programming of the Minimum S2/S3 parameter was dominated by nominal values in each zone. The details of zone-based programming are in Table S3.

## FIGURES

**Figure S1:**
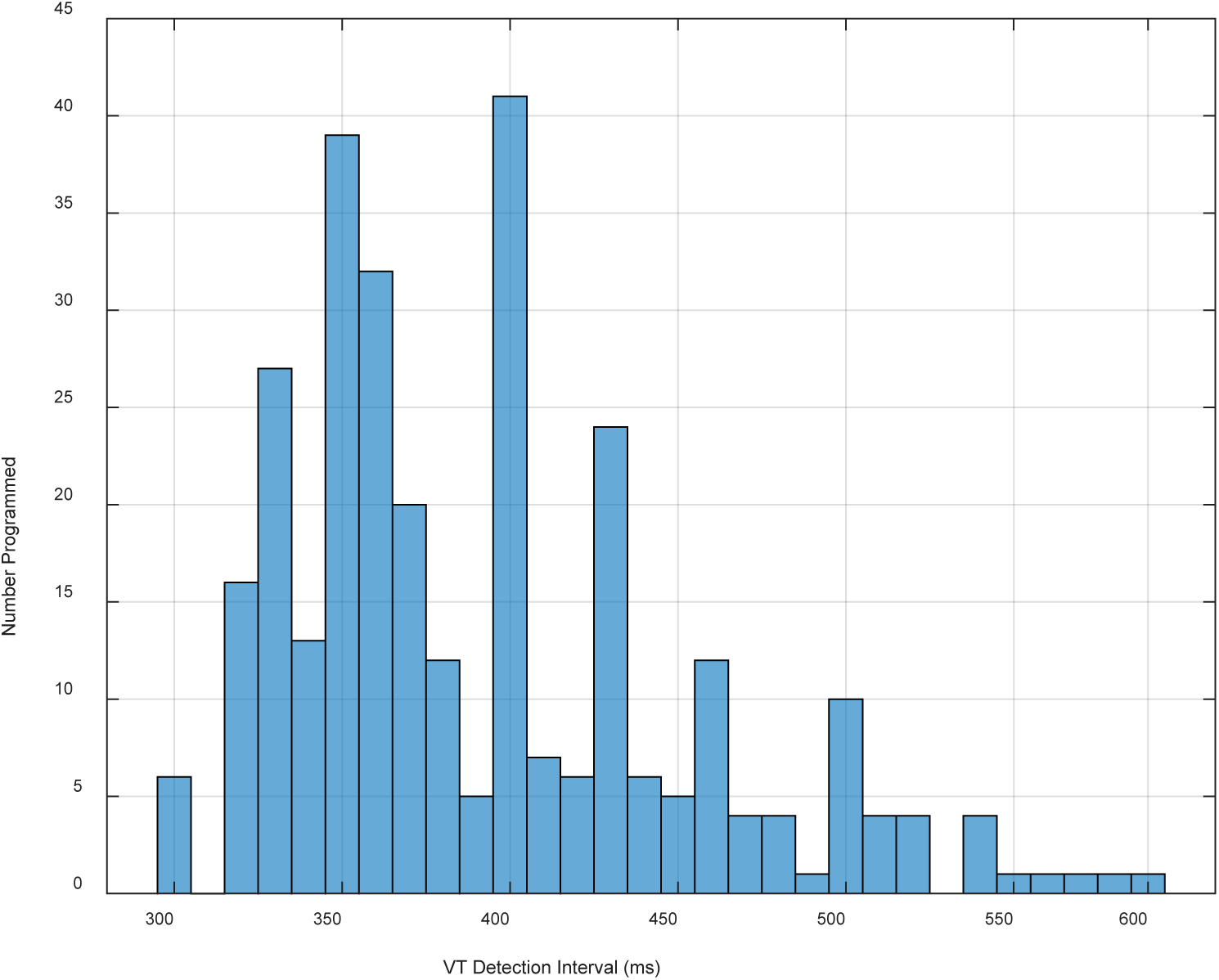
Histogram of the programmed values of the VT Detection Interval in the devices with MVT episodes.

**Figure S2:**
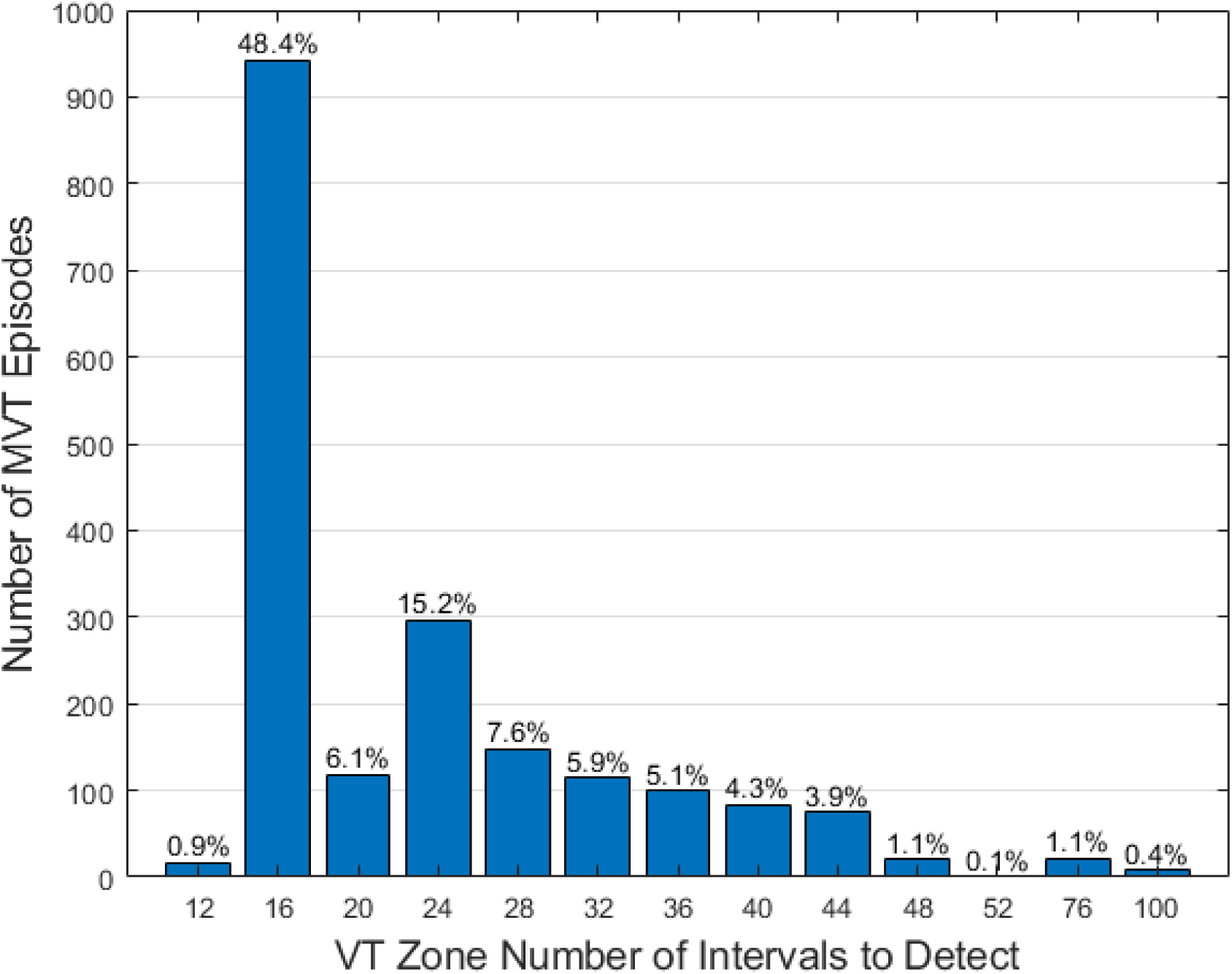
Histogram of the programmed VT Number of Intervals to Detect for MVT episodes. The percentage on the top of each bar is the proportion of the total number of MVT episodes at that initial detection setting in the VT zone (including FVT via VT). The nominal value is 16.

**Figure S3:**
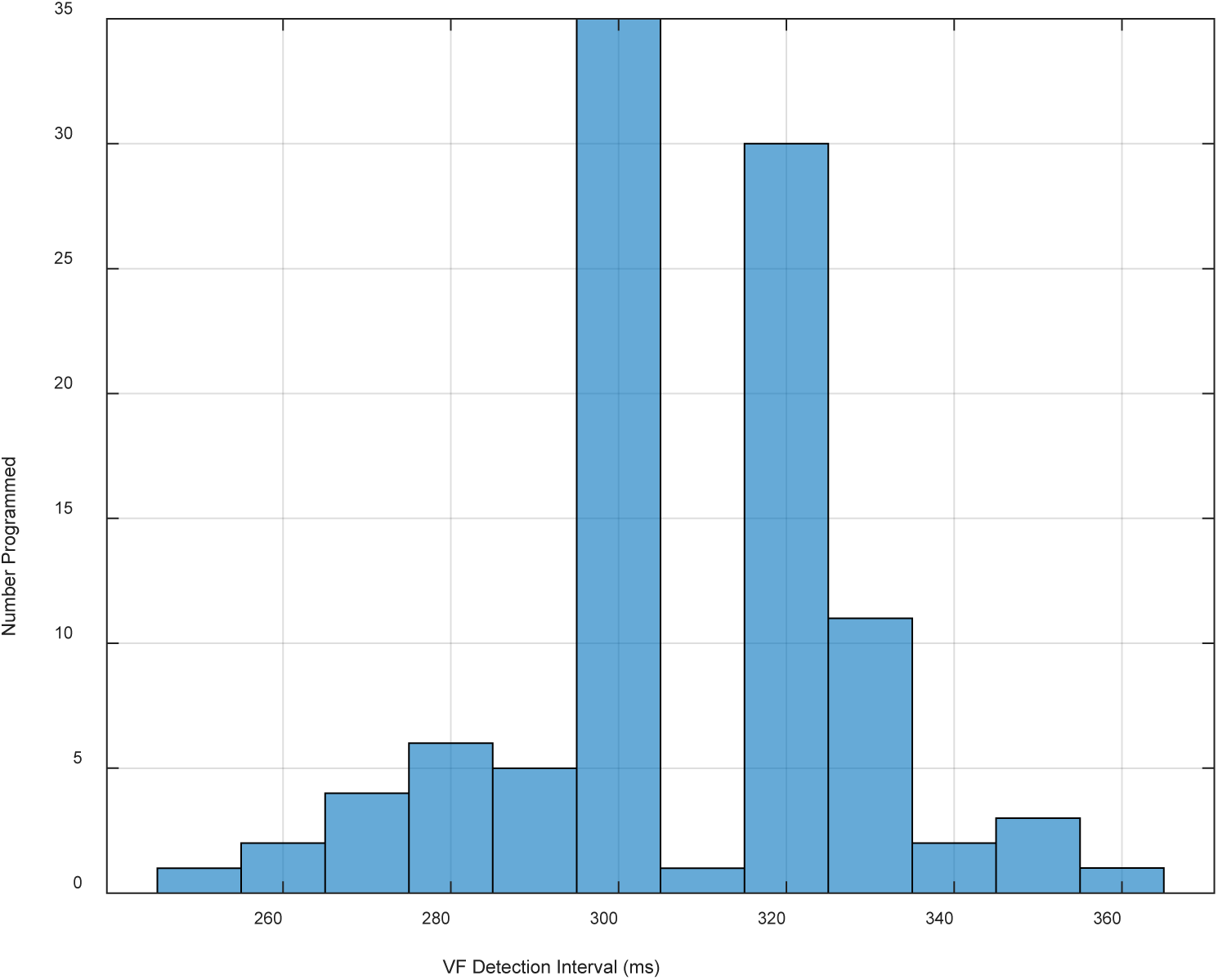
Histogram of the number of configurations programmed with a given VF Detection Interval in devices with MVT episodes. The nominal value of 320ms is the second most frequently used value.

**Figure S4:**
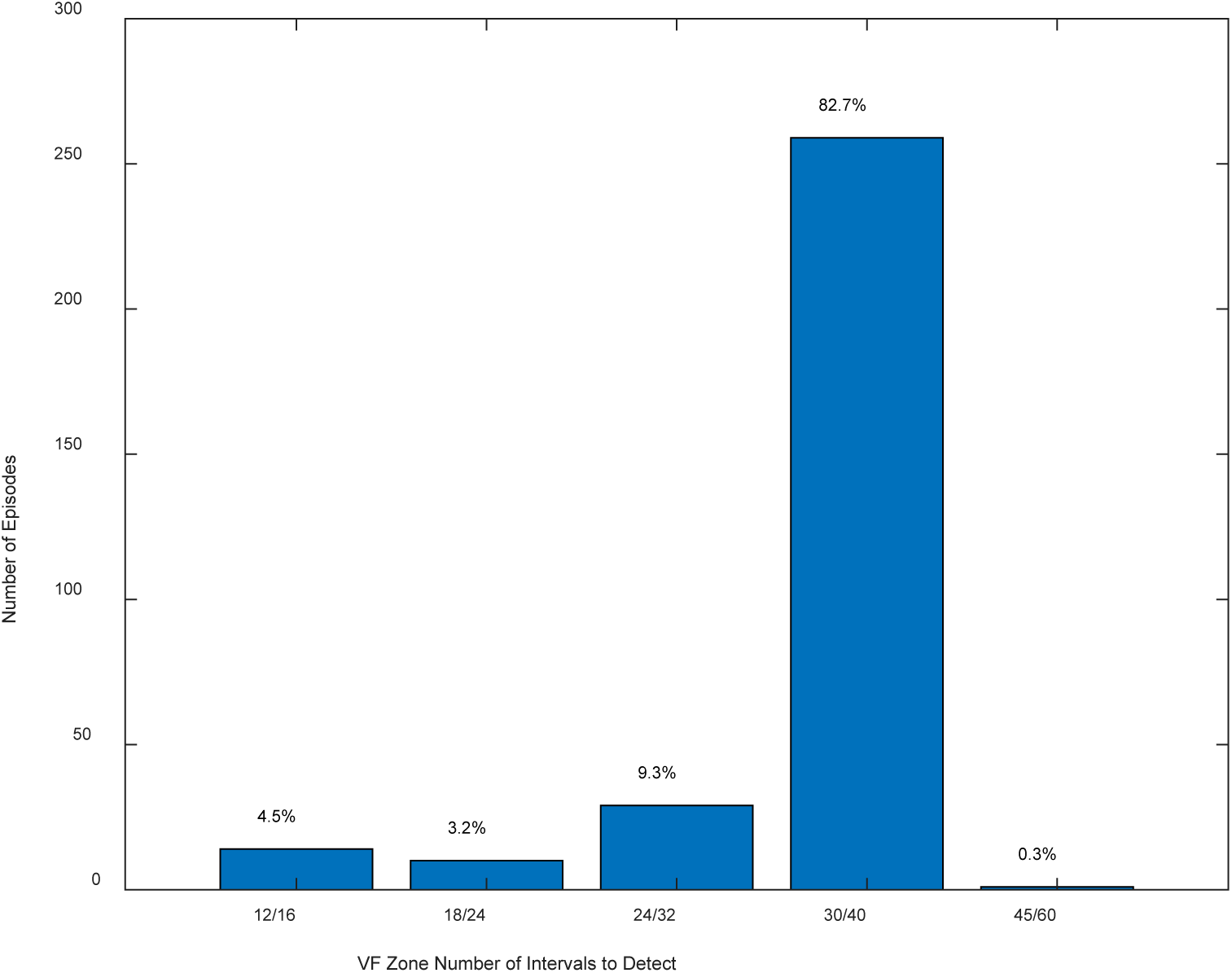
Histogram of the programmed VF Number of Intervals to Detect for MVT episodes. The percentage on the top of each bar is the proportion of the total number of MVT episodes at that initial detection setting in the VF zone (including FVT via VF). The nominal value is 30/40.

## TABLES

**Table S1:** Mapping of indication for device to key terms.

| Assigned indication | Term in patient record |
| --- | --- |
| Secondary prevention | "SECONDARY PREVENTION - PT SHOCKED PREV" |
|  | "PRIOR SUDDEN CARDIAC ARREST/VF" |
|  | "HISTORY OF SPONTANEOUS SUSTAINED VT" |
|  | "SYNCOPE WITH INDUCIBLE VT/VF" |
| Primary prevention | "PRIMARY PREVENTION - PT NEVER SHOCKED" |
|  | "DILATED CARDIOMYOPATHY" |
|  | "CARDIOMYOPATHY" |
|  | "HOCM (HYPERTROPHIC OBSTR CARDIOMYOPATHY)" |
|  | "LOW EF, DYSSYNCHRONY (WIDE QRS)" |
|  | "LOW EF, NO HISTORY VT/VF (SCD-HEFT)" |
|  | "NYHA CLASS III" |
| Other indication | "OTHER" |
|  | "SYNCOPE" |

**Table S2:** Programming of FVT Sub-zone Boundaries.

| **FVT via VF - minimum interval**<br>**for FVT classification** | **Number of patients**<br>**with via VF interval** | **FVT via VT –**<br>**maximum interval**<br>**for VT classification** | **Number of patients**<br>**with via VT interval** |
| --- | --- | --- | --- |
| 200 ms | 1 | 300 ms | 1 |
| 240 ms (nominal via VF) | 15 (44%) | 320 ms | 1 |
| 250 ms | 2 | 330 ms | 2 |
| 260 ms | 4 | 350 ms | 1 |
| 270 ms | 5 | 410 ms | 1 |
| 280 ms | 4 | 430 ms | 1 |
| 290 ms | 3 |  |  |

**Table S3:**
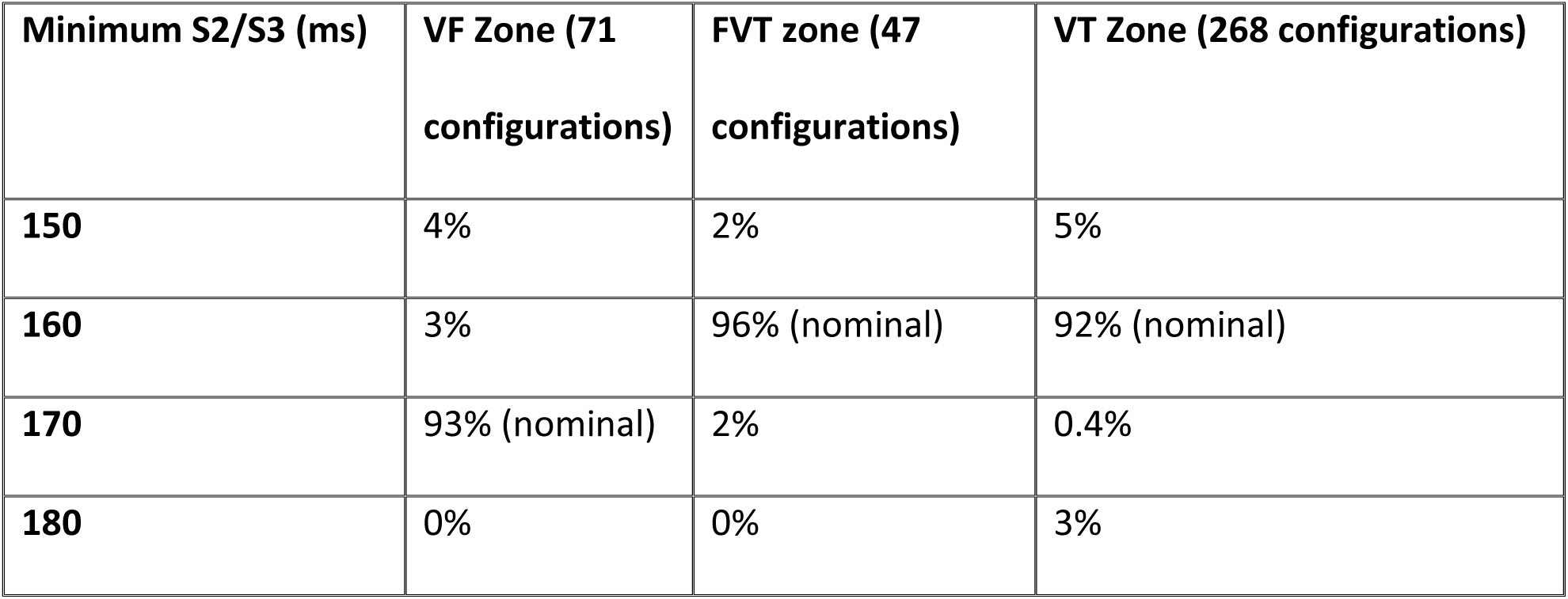
Programming of Minimum S2/S3 for Patients with Treated MVT in a Zone.

| **Minimum S2/S3 (ms)** | **VF Zone (71 configurations)** | **FVT zone (47 configurations)** | **VT Zone (268 configurations)** |
| --- | --- | --- | --- |
| **150** | 4% | 2% | 5% |
| **160** | 3% | 96% (nominal) | 92% (nominal) |
| **170** | 93% (nominal) | 2% | 0.4% |
| **180** | 0% | 0% | 3% |

---

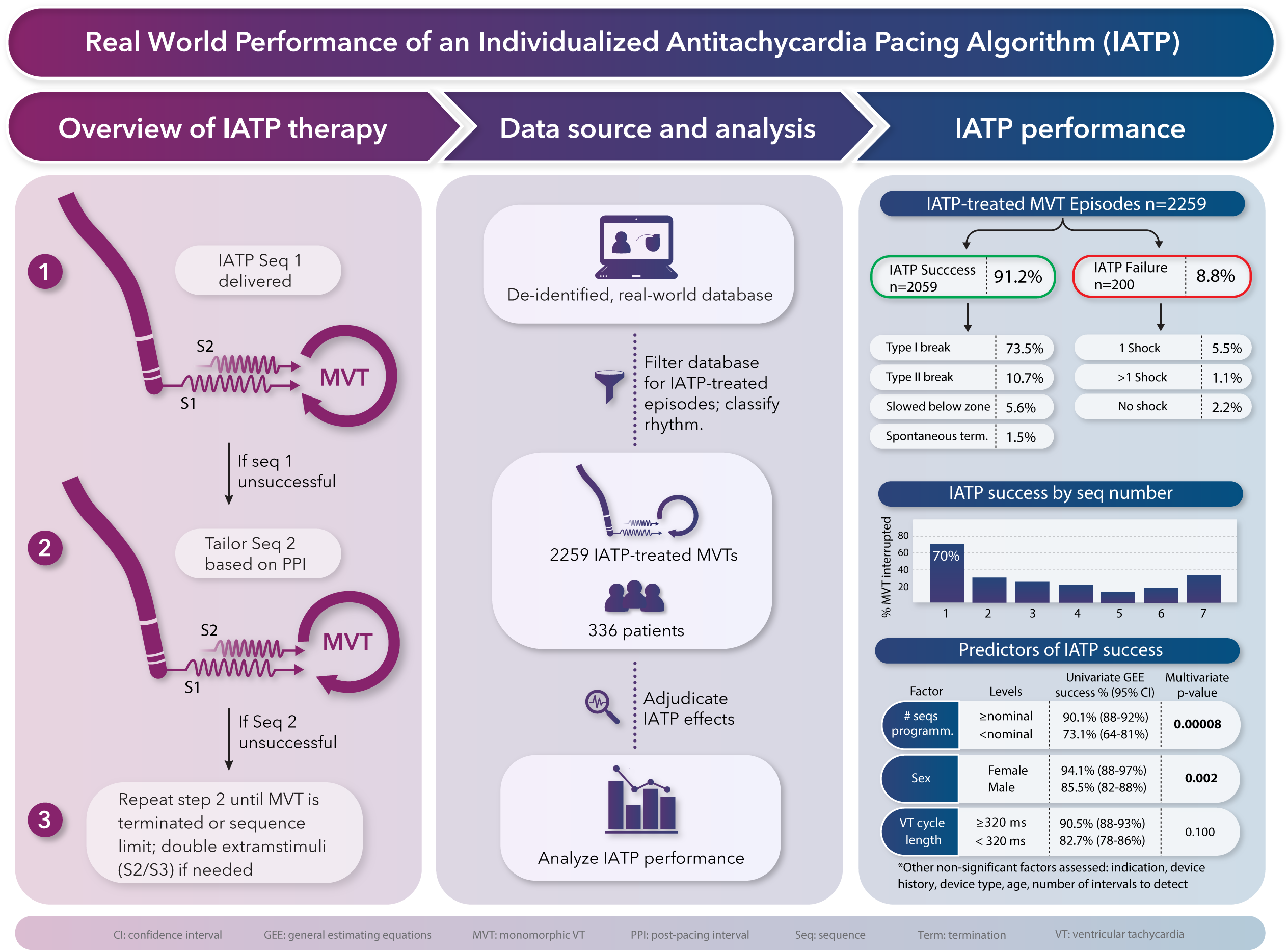

